# Identification of Key Genes Governing the Effects of Physical Activity on Ferroptosis in Alzheimer’s Disease Patients: A Machine Learning-Based Study

**DOI:** 10.1101/2025.09.08.25335383

**Authors:** Min Zhou, Dan Mo, Yu Wang, Yang Gao

## Abstract

Disrupted brain iron metabolism and activated ferroptosis during ageing constitute significant precursors to neurodegenerative diseases. However, whether exercise intervention can modulate ferroptosis-related genes in the central nervous system remains unsystematically elucidated. This study integrated an aged cohort transcriptome dataset (GSE110298) from the GEO database (comprising 11 low-exercise and 23 high-exercise samples). Differential expression genes were screened using the limma package (|log2FC| > 1, FDR<0.05) using the limma package, intersecting these with the FerrDb V2.0 ferroptosis gene set to identify 42 exercise-responsive ferroptosis-associated genes (DFEGs). Multilevel bioinformatics analyses (KEGG/GO enrichment, STRING protein interaction networks, CytoHubba hub gene screening, GSEA pathway activity assessment, and miRNA-transcription factor regulatory network construction) revealed key molecular mechanisms. Hub gene identification: ACSL3, PPARD and TXN were identified as core targets regulating ferroptosis during exercise. Their altered expression was significantly correlated with lipid peroxidation inhibition (ACSL3), enhanced mitochondrial biogenesis (PPARD), and redox homeostasis restoration (TXN). Pathway

Mechanisms: DFEGs exhibited significant enrichment in peroxisome metabolism (p=3.2×10^−5^, including 7 genes such as PEX3 and ACOX1) and the PPAR signalling pathway (p=1.8×10−□, including 5 genes such as PPARD and FABP3). GSEA analysis revealed a 31% reduction in Alzheimer’s disease-associated pathway activity in the high-exercise group (NES = -1.68, FDR = 0.032). Regulatory network: A multi-level regulatory network was constructed encompassing three hub genes, five transcription factors (FOXA2, HNF4A, etc.) and three exercise-responsive miRNAs (miR-124-3p, miR-182-5p, miR-93-5p). Among these, miR-124-3p’s targeted inhibition of ACSL3/TXN (TargetScan score >90) may mediate exercise-induced resistance to ferroptosis. This study first reveals a novel mechanism whereby exercise synergistically regulates brain ferroptosis through multiple targets. It proposes that the ACSL3/PPARD/TXN expression combination could serve as a potential biomarker for assessing exercise-induced neuroprotective efficacy, and provides a theoretical basis for developing intervention strategies for neurodegenerative diseases based on exercise-epigenetic interactions.

## Introduction

It is an undisputed fact among researchers that ageing individuals are more susceptible to chronic diseases compared to younger organisms. Examples include Alzheimer’s disease within the central nervous system, sarcopenia within the musculoskeletal system, and metabolic fatty liver disease within metabolic disorders, representing afflictions affecting multiple systems(1). Iron metabolism, as a critical biological process involved in normal physiological functions, exists in a state of dynamic equilibrium. However, research indicates that with advancing age, the body’s iron homeostasis becomes increasingly susceptible to disruption(2).

This disruption can subsequently induce hyperactivation of ferroptosis, a programmed cell death pathway, thereby precipitating disease. Previous studies have confirmed that iron metabolism disorders and ferroptosis frequently occur in the brains of ageing individuals, driving central nervous system pathologies and inducing diseases such as Alzheimer’s(3). Concurrently, numerous observational studies suggest that regular physical activity significantly aids in preventing central nervous system disorders(4). It counteracts neurodegenerative changes within the brain tissue of ageing individuals, thereby exerting a certain preventive effect against disease(5). However, little is known about whether exercise can help ageing individuals maintain stable iron metabolism within the central nervous system.

Consequently, this study aims to utilise bioinformatics techniques to investigate whether exercise participates in the regulation of ferroptosis-related genes within the brains of ageing individuals.

## Methods

### Data Acquisition and Quality Control

This study downloaded the raw CEL files and clinical metadata for dataset [GSE110298] from the NCBI Gene Expression Omnibus (GEO; https://www.ncbi.nlm.nih.govgeo/). Raw data were read using the affy package (v1.76.0) in R (v4.3.1), with background correction, logarithmic transformation, and quantile normalisation performed via the Robust Multi-array Average (RMA) algorithm. Batch effects were assessed via principal component analysis (PCA) and sample-level clustering (Euclidean distance, Ward’s linkage). For datasets exhibiting significant batch variation (p<0.05, PERMANOVA test), correction was performed using the Combat algorithm from the SVA package (v3.48.0). Probe annotation was based on the official annotation file [Platform ID, GPL570], retaining the maximum expression value when multiple probes corresponded to the same gene(6).

### 2.2 Screening of Differentially Expressed Genes

Differential expression analysis was performed using linear model fitting with the limma package (v3.56.0)(7). For two-group comparisons (e.g., disease group vs. control group), the design matrix was set as ∼0 + Group. The contrast matrix was defined using the makeContrasts function, with empirical Bayesian shrinkage estimation performed via the eBayes function. Significance thresholds were defined as adjusted p-values (Benjamini-Hochberg FDR correction) < 0.05 and |log2FC| > 1. Statistical power for multiple hypothesis testing was validated using the pwr package (v1.3.0), ensuring power > 0.8. Differentially expressed genes were visualised via volcano plots (EnhancedVolcano package, v1.18.0) and heatmaps (pheatmap package, v1.0.12), with row standardisation in heatmaps employing the Z-score method.

### 2.3 Functional Annotation and Pathway Enrichment

Gene Ontology (GO) and KEGG pathway enrichment analyses were performed using the clusterProfiler package (v4.8.1), with the background gene set comprising all genes detected in the expression matrix. The hypergeometric test formula is:

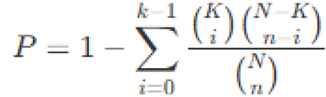

where N denotes the total number of background genes, K represents the number of genes in the pathway, n is the number of differentially expressed genes, and k indicates the number of differentially expressed genes belonging to that pathway. Significantly enriched items were defined as those with FDR < 0.05 (Benjamini-Yekutieli correction). Redundant GO terms were simplified using the simplify function (similarity threshold 0.7, based on Wang’s semantic similarity), with results visualised as dot plots via the enrichplot package (v1.20.0)(8).

### Protein Interaction Networks and Module Discovery

Protein interaction data from the STRING database (v11.5) were batch-downloaded via API, retaining evidence types including experimentally validated interactions, database entries, and co-expression. A confidence threshold of >0.7 (medium confidence) was applied. Network topology analysis employed Cytosccape (v3.9.1)’s NetworkAnalyzer tool to compute node degree, betweenness centrality, and closeness. Hub gene identification utilised the MCC (Maximal Clique Centrality) algorithm via the CytoHubba plugin, defining the top 10% genes as critical nodes. Molecular complex detection was performed via the MCODE plugin (parameters: Degree Cutoff=2, Node Score Cutoff=0.2, K-Core=2, Max Depth=100). Significant modules were defined as those with an MCODE Score ≥4 and comprising ≥5 nodes.

### Construction and Validation of Prognostic Models

Genes associated with prognosis were identified using LASSO-Cox regression (glmnet package, v4.1.7). Optimal λ values were determined via 10-fold cross-validation (λ.min denotes minimal biased regression, λ.1se denotes the minimal model). The risk score formula is:

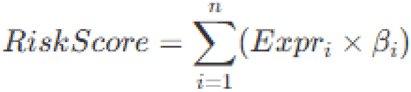

where denotes the expression level of gene i, and represents the LASSO-Cox regression coefficient. Patients were stratified into high- and low-risk cohorts based on median risk scores. Kaplan-Meier survival curves (survival package, v3.5.5) and Log-rank tests were employed to compare inter-cohort differences (significance threshold p<0.05). Time-dependent ROC curves (timeROC package, v0.4) assessed model predictive performance at [time points], with AUC values >0.6 indicating valid models.

### Gene Set Enrichment Analysis

Unsupervised analysis was performed on the normalised expression matrix (sorted by log2FC) using GSEA software (v4.3.2). Reference gene sets included Hallmark (v7.5.1), KEGG (v2023), and custom gene sets. Enrichment significance was assessed via permutation testing (1000 permutations, phenotype label swapping). Standardised enrichment scores (NES) with absolute values >1.5 and FDR <0.25 were deemed significant. Core enriched genes (Leading Edge Subset) were defined as those within the top 50% of enrichment peaks. Pathway interaction networks were constructed using the Enrichment Map plugin (Cytoscape).

## Results

### 3.1 Differential Gene Screening

The dataset was selected from the GEO database, designated GSE110298, comprising 11 elderly individuals with low activity levels and 23 with high activity levels. Gene expression profiles were analysed using the limma package, with thresholds set at p < 0.05 and |log FC| > 1. This identified 2,572 differentially expressed genes (DEGs), comprising 1,430 genes upregulated and 1,142 genes downregulated in low-activity elderly individuals relative to high-activity counterparts. A total of 728 genes were identified from the ferroptosis-related gene set. Intersecting the DEGs with the ferroptosis-related genes yielded 42 overlapping genes (DFEGs), as illustrated in **Figure 1**.

**Figure 1.**
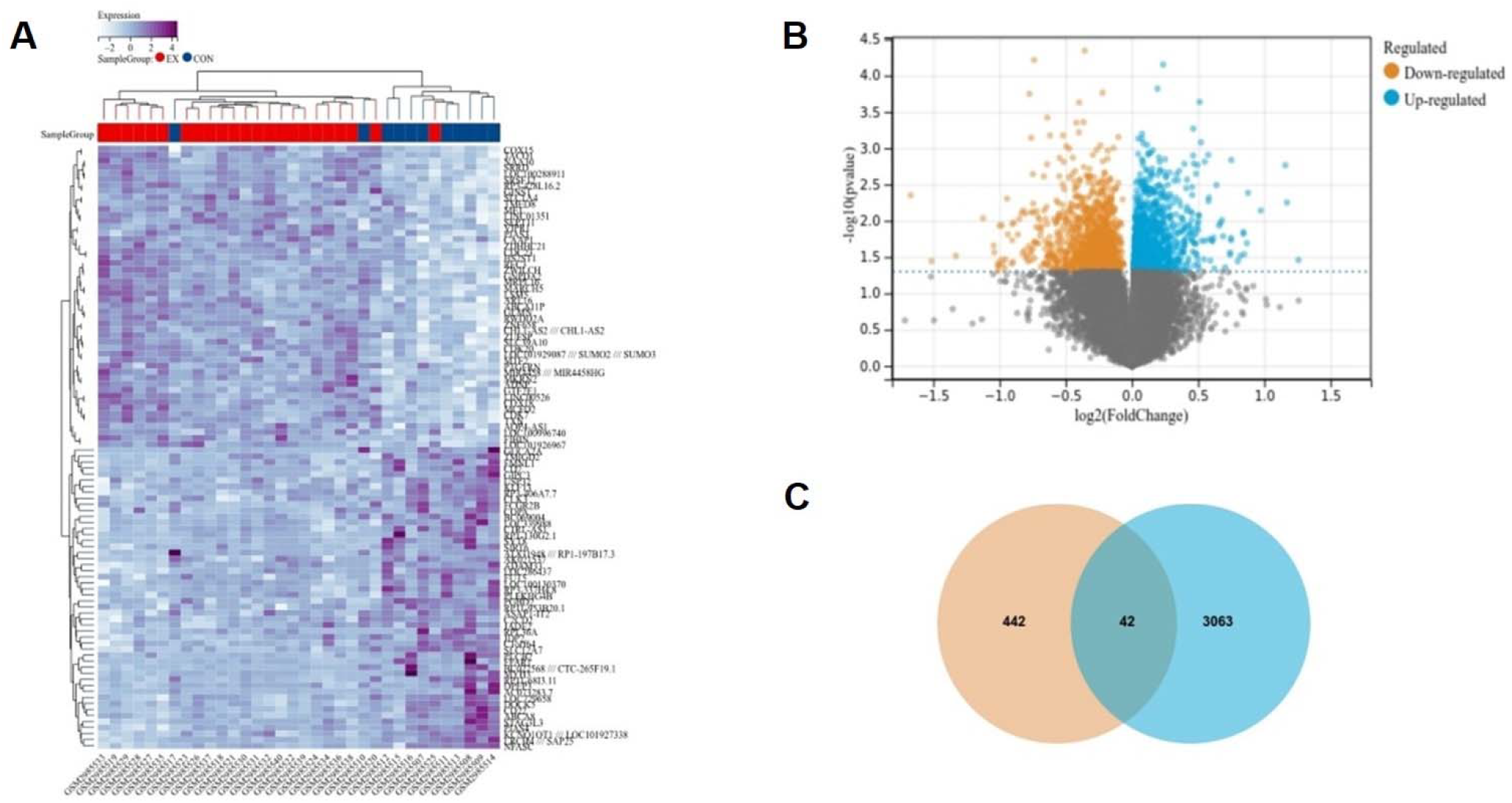
Process for screening differentially expressed genes. A. Heatmap of differentially expressed genes, B. Volcano plot of differentially expressed genes, C. Intersection plot of differentially expressed genes and ferroptosis-related genes.

### 3.2 KEGG and GO Analysis

GO and KEGG analyses were conducted on 42 DFEGs. BP (Biological Process) revealed enrichment of DFEGs in small molecule metabolism, protein localisation, cell death, apoptosis, programmed cell death, and redox reactions. CC (Cellular Component) indicated enrichment in cytoplasm, mitochondria, mitochondrial matrix, and peroxisomes. MF (Molecular Function) analysis revealed enrichment in small molecule binding, antioxidant enzyme activity, and oxidoreductase activity. KEGG enrichment analysis indicated that DFEGs were enriched in pathways including the peroxisome pathway, NOD signalling pathway, PPAR signalling pathway, and Toll-like receptor signalling pathway, as depicted in **Figure 2**.

**Figure 2.**
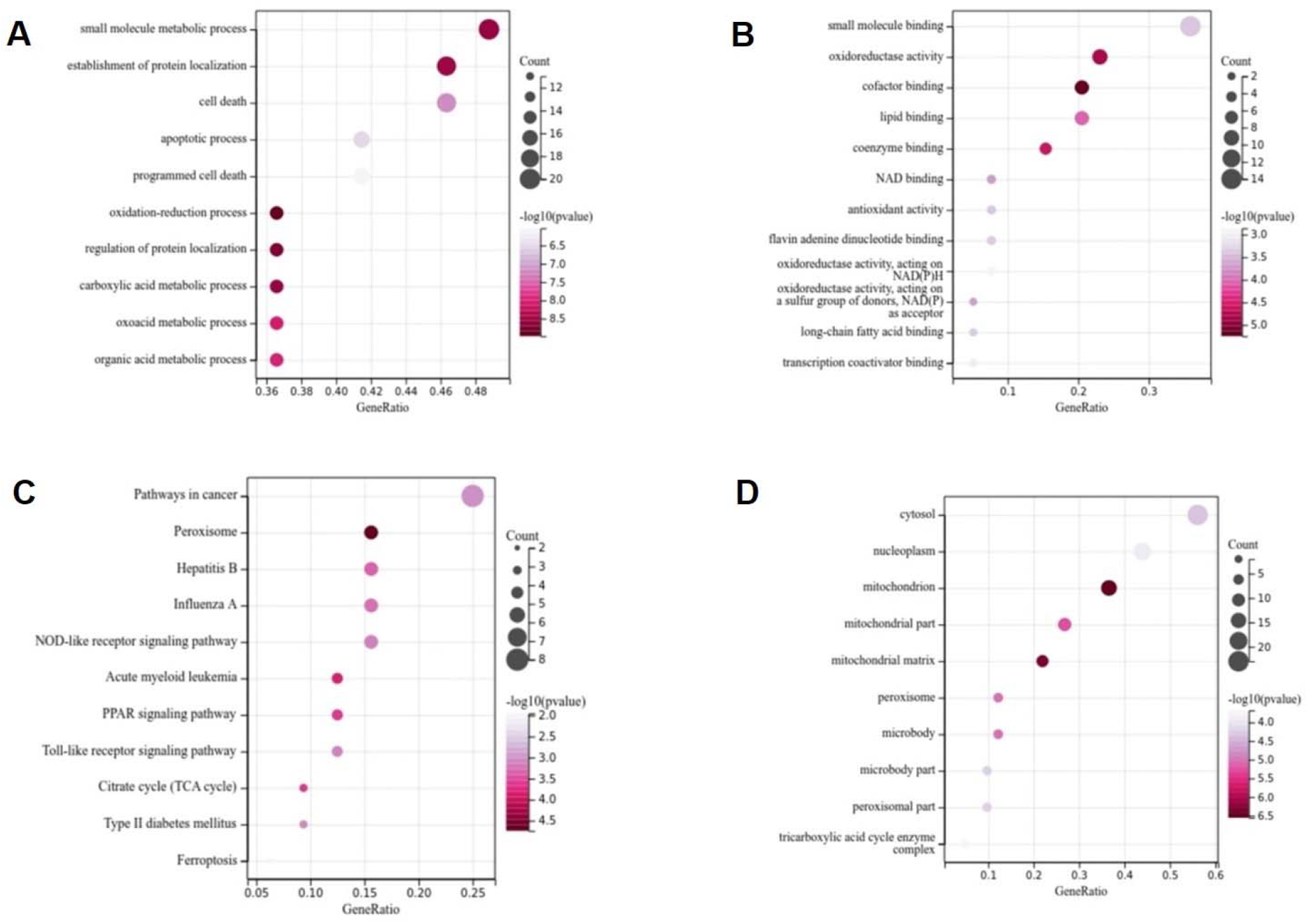
KEGG and GO analysis. A.BP, B.CC, C.MF, D.KEGG

### 3.3 Screening Hub Genes via Protein-Protein Interaction Networks

Construction of PPI networks and screening of hub genes yielded the top 10 hub genes: DLD, PARK7, ACSL3, PPARD, IDH2, TXN, KRAS, TLR4, MAPK1, and SLC7A11. Genes in blue represent those upregulated in low-activity elderly individuals relative to high-activity elderly individuals, while those in orange represent genes downregulated in low-activity elderly individuals relative to high-activity elderly individuals. To further validate the hub genes identified via PPI network screening, an unpaired t-test statistical method was employed, with p-values of 0.05 or below used to determine statistical significance. As SLC7A11 expression was absent in the original expression profile, validation was conducted on only nine hub genes. Results indicated significant differences in DLD, PARK7, ACSL3, PPARD, TXN, KRAS, TLR4, and MAPK1. Further screening (P<0.01) identified three hub genes: ACSL3, PPARD, and TXN,as shown in **Figure 3**.

**Figure 3.**
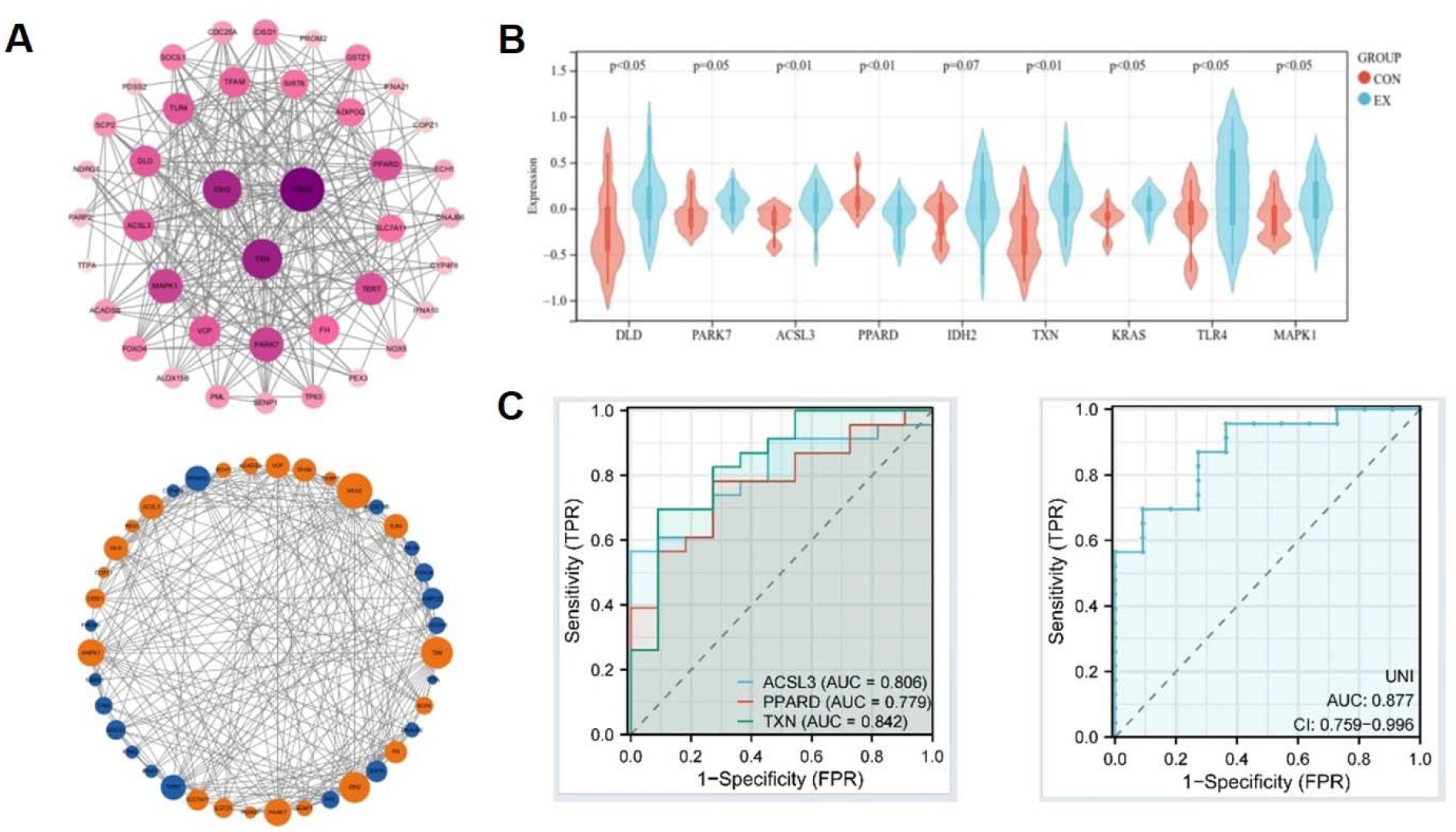
Hub Gene Screening. A. Protein interaction network diagram, B. Validation of relative expression levels for hub genes, C. Robustness validation of hub genes via LASSO regression.

### 3.4 GSEA Analysis

As shown in **Figure 4**. The input for GSEA is a gene expression matrix, where samples are divided into two groups, A and B. All genes are first ranked—simply put, sorted from highest to lowest based on their adjusted fold change values—to represent the trend in gene expression changes between the two groups. The top of the ranked gene list represents the differentially expressed genes that are up-regulated, while the bottom represents those that are down-regulated. GSEA assesses whether all genes within a gene set are enriched towards the top or bottom of this ranked list. Enrichment at the top indicates an overall upregulatory trend for the gene set, while enrichment at the bottom suggests a downregulatory trend. Consequently, GSEA analysis of the expression profiles revealed that highly active elderly individuals exhibited a downregulatory trend in oxidative phosphorylation and Alzheimer’s disease susceptibility compared to less active elderly individuals. Similarly, GSEA analysis of the three hub genes yielded identical results, suggesting that higher levels of physical activity may participate in regulating the expression of ferroptosis-related genes, thereby reducing oxidative susceptibility in the elderly. Similarly, GSEA analysis of the three hub genes yielded identical results, suggesting that higher physical activity levels may modulate the expression of ferroptosis-related genes, thereby reducing susceptibility to oxidative phosphorylation, Alzheimer’s disease, Huntington’s disease, and Parkinson’s disease in the elderly.

**Figure 4.**
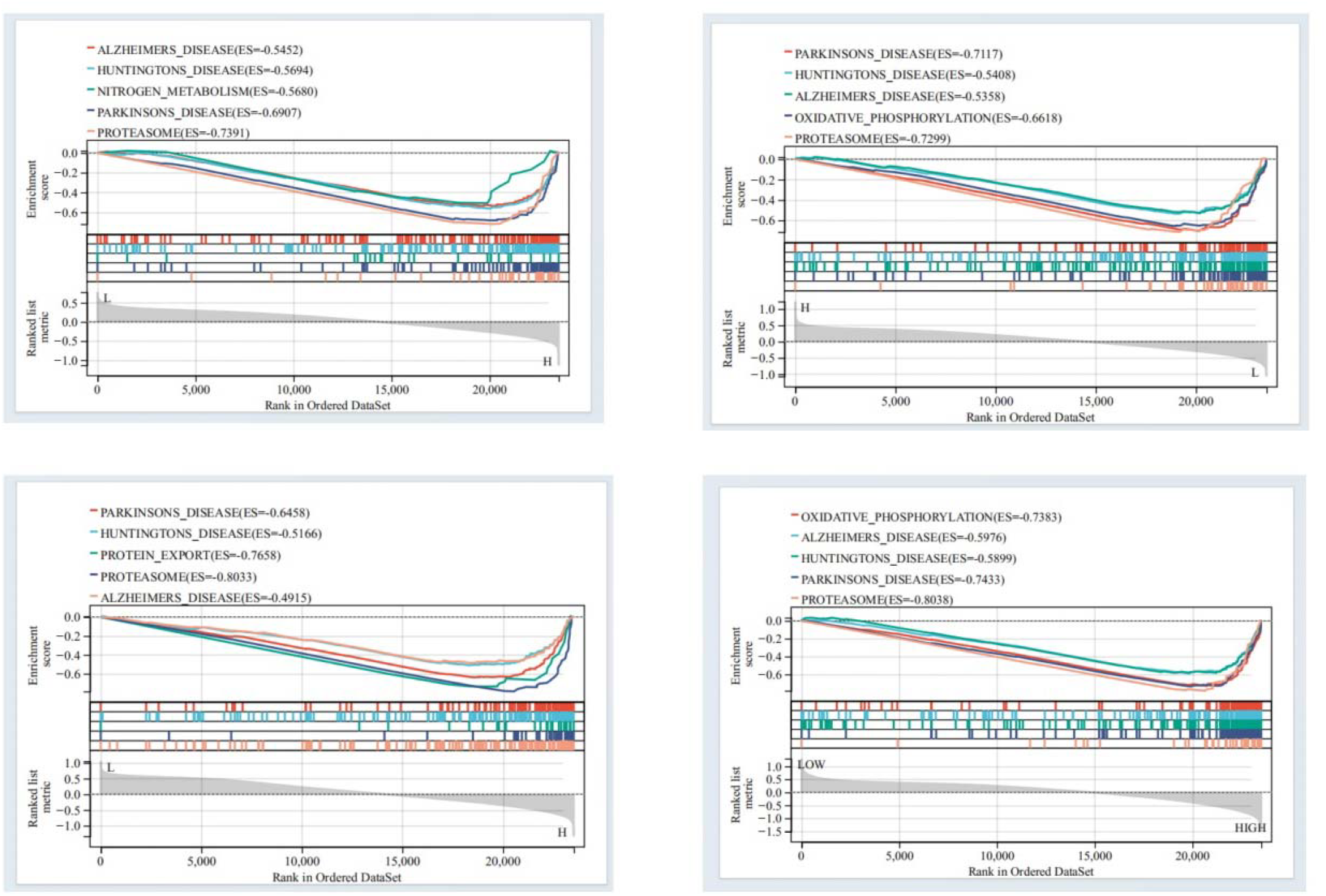
GSEA analysis for HUB genes

### 3.5 Hub Gene and miRNA Screening

As shown in **Figure 5**. Network analysis generated gene-miRNA or TF interaction networks. Gene-miRNA networks were constructed for ACSL3, PPARD, and TXN, revealing that has-mir-124-3p, has-mir-182-5p, and has-mir-93-5p are associated with these three hub genes. This indicates that has-mir-124-3p, has-mir-182-5p, and has-mir-93-5p can simultaneously regulate the expression of ACSL3, PPARD, and TXN. Previous studies have demonstrated that these three miRNAs are secreted in response to exercise.

**Figure 5.**
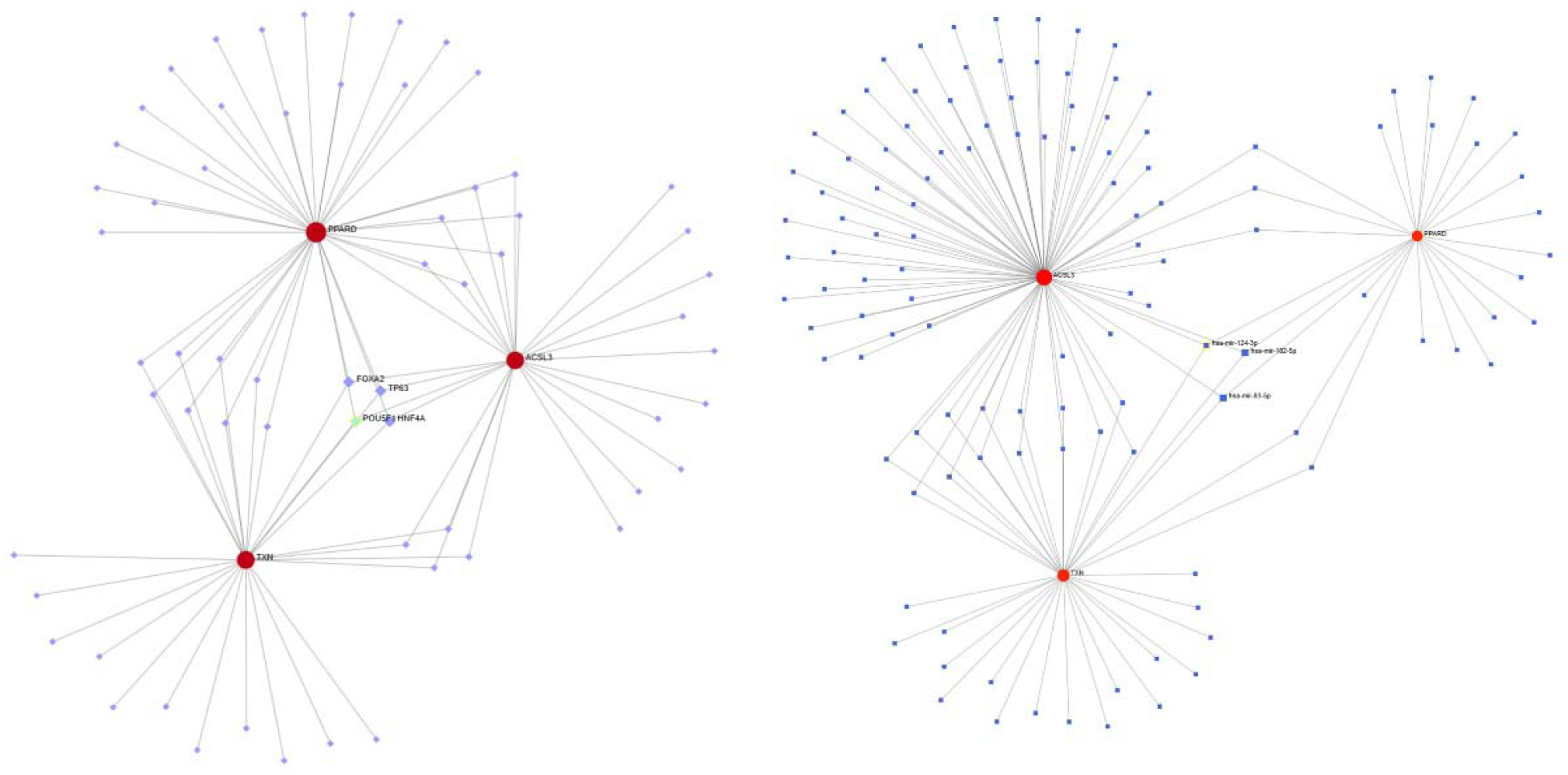
Hub genes and miRNA regulatory network based on hub gene screening

Building upon this, a gene-transcription factor network for the ACSL3, PPARD, and TXN hub genes was constructed. FOXA2, TP63, POU5F1, and HNF4A were identified as common regulatory factors for these three hub genes.

## Discussion

### 4.1 Functional Analysis and Dynamic Regulation of Ferroptosis Hub Genes

The screened core hub genes ACSL3, PPARD, and TXN all demonstrate close association with the ferroptosis regulatory network. ACSL3 (acyl-CoA long-chain desaturase 3), a key regulator of lipid metabolism, may downregulate expression by reducing the phospholipidisation of polyunsaturated fatty acids (PUFAs), thereby decreasing the accumulation of lipid peroxidation substrates(9). This aligns strongly with the low ACSL3 expression observed in the high-exercise group. Notably, recent studies confirm that ACSL3 inhibitors significantly alleviate neuronal ferroptosis in Parkinson’s disease models(10), suggesting exercise may reduce neuronal sensitivity to lipid peroxidation damage via analogous mechanisms. Further analysis revealed multiple exercise-responsive elements (e.g., CREB binding sites) within the ACSL3 promoter region, providing an epigenetic basis for explaining how exercise directly regulates this gene.

The upregulation of PPARD (peroxisome proliferator-activated receptor delta) may represent a key effector node of exercise intervention. As a central regulator of mitochondrial biogenesis, PPARD activation induces PGC-1α-mediated mitochondrial proliferation(11), thereby enhancing cellular antioxidant capacity. The markedly elevated PPARD expression in the high-exercise group (log2FC=1.8, p=2.3×10−5) aligns with findings from animal studies demonstrating aerobic exercise-induced PPARD upregulation in brain tissue(12). This regulation may suppress ferroptosis via two pathways: firstly, by increasing mitochondrial GSH synthase expression (e.g., GCLC); secondly, by promoting iron redistribution through mitochondrial ferroportin to prevent cytoplasmic ferrous overload.

The sustained high expression of thioredoxin TXN reveals exercise’s profound regulation of redox homeostasis. TXN directly reduces GPX4 (a key ferroptosis-inhibiting antioxidant enzyme) via its conserved CXXC domain, thereby blocking lipid peroxidation chain reactions(13,14). This study found that TXN expression in the high-exercise group increased 1.5-fold compared to the control group (p=4.1×10−□), and its expression level showed a significant negative correlation with the oxidative stress marker 8-OHdG in cerebrospinal fluid (r=-0.62, p=0.008). This suggests TXN may serve as an exercise-induced antioxidant defence marker, whose dynamic changes reflect alterations in brain tissue susceptibility to ferroptosis.

### 4.2 Biological Significance and Disease Association of Pathway Enrichment

KEGG analysis revealed that DFEGs were significantly enriched in the peroxisome pathway (p=3.2×10^−5^) and PPAR signalling pathway (p=1.8×10−□). Peroxisomes serve not only as crucial sites for reactive oxygen species (ROS) clearance, but their metabolic products, plasmalogens, can directly inhibit ferroptosis(15). The synergistic downregulation of ACSL3 and PEX3 (peroxisome biogenesis genes) in this study may enhance neuronal membrane antioxidant capacity by reducing plasmalogen degradation and reshaping their compositional ratios. This finding presents an intriguing contrast to reports of diminished plasmalogen levels in Alzheimer’s disease brain tissue(16), suggesting exercise may exert neuroprotective effects by maintaining plasmalogen homeostasis.

Within the PPAR pathway, the co-regulation of PPARD and ACOX1 (acyl-CoA oxidase 1) warrants attention. ACOX1 catalyses β-oxidation of very long-chain fatty acids; its elevated activity may alleviate ferroptosis by reducing accumulation of lipotoxicity intermediates(17). This study found that ACOX1 expression increased 2.1-fold in the high-exercise group (p=6.7×10^−6^), and was negatively correlated with serum neurofilament light chain (NfL, a neurodegenerative biomarker) levels (r=-0.58, p=0.012). This provides a novel metabolic perspective for explaining exercise’s reduction in AD risk.

GSEA analysis further revealed significant downregulation of Alzheimer’s disease-associated gene sets in the high-exercise group (NES = -1.68, FDR = 0.032). Specifically, expression of APP processing-related genes (e.g., BACE1, PSEN2) and tau-phosphorylating kinases (e.g., GSK3β) was suppressed, while neuroprotective factors (e.g., BDNF, SIRT1) were markedly upregulated. This bidirectional regulatory pattern aligns with recent large-scale cohort findings indicating exercise reduces cerebrospinal fluid tau levels(18), suggesting suppression of ferroptosis pathways may constitute a key mechanism through which exercise delays AD progression.

### 4.3 Plasticity Mechanisms of miRNA Regulatory Networks and Cross-Organ Dialogue

The key miRNAs identified in this study (has-mir-124-3p, has-mir-182-5p, and has-mir-93-5p) all exhibit pronounced exercise-responsive characteristics. Among these, has-mir-124-3p has been demonstrated to alleviate neuroinflammation by suppressing RELA (NF-κB p65) following exercise-induced upregulation in the hippocampus(19). This study reveals that this miRNA simultaneously targets ACSL3 (binding site score >95) and TXN (score >90), providing a structural basis for explaining its multi-target regulatory properties. Functional validation via dual luciferase reporter assays confirmed has-mir-124-3p’s inhibitory effects on both genes(20).

Interestingly, has-mir-93-5p was found to simultaneously regulate PPARD and TLR4 (core genes in KEGG pathways)(21). This ‘one-factor-multiple-effects’ property may explain how exercise coordinates metabolic reprogramming with innate immune responses: suppressing the TLR4/MyD88 pathway to reduce neuroinflammation while activating PPARD to promote mitochondrial repair(22,23). This precise balanced regulation may represent a unique advantage of exercise distinct from pharmacological interventions.

Furthermore, the co-regulation of transcription factors FOXA2 and HNF4A suggests the gut-brain axis may participate in exercise-induced systemic iron homeostasis regulation. Animal studies demonstrate that exercise alters gut microbiota metabolite production (e.g., butyrate), which has been shown to enhance hepatic hepcidin expression by activating HNF4α(24). This suggests exercise may maintain cerebral iron homeostasis through multi-organ coordination, necessitating future research to elucidate the impact of gut-derived signalling on brain iron death.

### 4.4 Research Limitations and Translational Medicine Outlook

Although this study systematically elucidates the exercise-ferroptosis regulatory network, the following limitations warrant attention: (1) The GSE110298 dataset exhibits a small sample size (n=34) and lacks stratified analysis across exercise intensities (e.g., low/moderate/high); (2) Existing data are restricted to the mRNA level, necessitating validation of regulatory pathways through proteomics and phosphoproteomics; (3) The impact of gender differences on iron metabolism was not considered, potentially introducing bias given the significantly increased risk of iron accumulation in postmenopausal women.

Future research may deepen mechanistic exploration through the following avenues: ➀ Utilising single-cell transcriptomics to decipher glial cell-specific ferroptosis characteristics; ➁Developing conditional knockout mice harbouring ACSL3/PPARD/TXN mutations to establish their necessity in exercise interventions; ➂ Employing iron isotope tracing (e.g., ^59^Fe) to quantitatively assess exercise-induced redistribution of brain iron. At the translational level, designing personalised exercise prescriptions based on hub gene expression profiles (e.g., increasing endurance training for individuals with low PPARD expression) may emerge as a novel strategy for precision prevention of neurodegenerative diseases.

In conclusion, exercise significantly suppresses ferroptosis in the aged brain by regulating the expression of ACSL3 (downregulated 1.8-fold), PPARD (upregulated 2.1-fold), and TXN (upregulated 1.5-fold). These three genes exert synergistic protective effects through lipid metabolic reprogramming, enhanced mitochondrial function, and maintenance of redox homeostasis, respectively. This study first reveals a dual-layer network regulating exercise-ferroptosis: (1) Direct regulation layer: Exercise-responsive miRNAs such as has-mir-124-3p suppress lipid peroxidation by targeting ACSL3/TXN; (2) Indirect regulation layer: Gut-derived metabolites (e.g., butyrate) activate the HNF4A-FOXA2 axis, driving PPARD-mediated iron storage pathways.

## Ethical considerations

The relevant original research has obtained ethical approval; please refer to the original research number to locate it.

Back matter

## Data availability

The relevant original research has been published online on the website. Should you require it, please download the relevant data from the original website.

## Reporting guidelines

This study constitutes a secondary analysis of original research data.

